# Multi-ancestral genome-wide study of chronic pain reveals widespread genetic correlations with mental and physical health traits

**DOI:** 10.1101/2025.09.29.25336894

**Authors:** Pamela N Romero Villela, Jared V Balbona, Nithya Sarabudla, Alexander S Hatoum, Simon Haroutounian, Arpana Agrawal, Emma C Johnson

## Abstract

Chronic pain (CP) is a debilitating condition that impacts an individual’s physical and mental health. CP is common, affecting around 12-40% of individuals across the world. In this study, we conducted a genome-wide association study (GWAS) of lifetime incidence of CP in the All of Us Research Program, across six different ancestries in the United States (total N_case_ = 137 043, total N_control_ = 154 038). We found one genome-wide significant locus in the cross-ancestry meta-analysis on chromosome 5 (lead SNP: rs1196962975, *p* = 4.53E-08). Additionally, one locus on chromosome 11 (lead SNP: rs5795684) reached genome-wide significance in the admixed genetic ancestry sample (*p* = 1.13E-08). Cross-ancestry genetic correlations ranged from 0.40 – 0.78. CP was genetically correlated with a range of psychiatric, physical health, and immune traits, including anxiety (r_g_ = 0.69, *p* = 1.82E-69), generalized addiction risk (r_g_ = 0.39, *p* = 1.98E-18), and increased serum C-reactive protein levels (r_g_ = 0.35, *p* = 5.28E-22). This study represents the largest cross-ancestral investigation of the genetics of lifetime incidence of chronic pain to date and demonstrates shared genetic effects between chronic pain and psychiatric disorders and other physical health conditions.

**Highlights:**

- 48.8% of individuals were recorded as having a lifetime incidence of chronic pain.
- One locus, rs5795684, was significant in individuals of admixed ancestry.
- One additional locus, rs1196962975, was significant in a cross-ancestral meta-analysis.
- Genetic associations were enriched in brain areas such as the nucleus accumbens and amygdala.
- Chronic pain was moderately to highly genetically correlated to other physical and psychiatric health traits.

## Introduction

Chronic pain (CP), defined as pain lasting 3 or more months^1^, is one of the most common health conditions in the United States (US)^2^. CP is debilitating, with negative effects on daily activities^3^ and overall mental health^4^. CP is highly complex; it may arise following another disease^5,6^, after a surgical procedure^7^, or may have unclear origins^8^. CP may also be classified in various ways, based on its location (e.g., back pain), cause (e.g., a physically strenuous event), or impacted body system (e.g., musculoskeletal pain vs. neuropathic pain)^9^. These different classification systems of CP showcase its multi-faceted and heterogeneous nature.

CP is moderately heritable; past studies^10^ estimated that the narrow-sense heritability (h^2^) for CP is ∼38.4%. Despite CP’s complexity and heterogeneity, genome-wide association studies (GWAS) have successfully identified genetic variants associated with CP. One study^11^ of multi-site CP in ∼380 000 British individuals found 39 independent loci associated with CP. Authors found that these loci were enriched in areas of the genome involved in the development of the nervous system, neurogenesis, synaptic plasticity, and cell death (i.e., apoptosis). Another study by Zorina-Lichtenwalter et al. ^12^ modeled the genetics of 24 types of CP conditions and found that there was significant shared genetic variance among the conditions, suggesting that there is substantial genetic and biological overlap even among different kinds of CP. These 24 CP conditions fell between two genetic factors: one for general pain and another for musculoskeletal pain, providing support for substantial genetic covariance across pain arising from different sources as well as for genetic factors unique to certain kinds of pain, such as musculoskeletal pain. Most recently, Toikumo et al.^13^ conducted the largest GWAS meta-analysis of CP and found 343 genomic risk loci in >1.2 million individuals; notably, this study only analyzed participants of European ancestry. All prior GWAS of generalized CP have only included individuals of European genetic ancestry. However, a GWAS^14^ of chronic back pain in the Million Veteran Program included over 553 000 individuals across three ancestry groups (African, European, and Admixed ancestry). This GWAS identified 87 genomic risk loci, 67 of which were novel. However, this study was limited to US veterans, and therefore, discovery of loci related to CP experienced by multiple civilian populations within the US remains lacking.

The All of Us Research Program (AoU)^15^ aims to spur novel biomedical research that improves health outcomes for all individuals. As part of this mission, AoU is one of the most diverse biobanks in the world, in terms of ancestry, age, socioeconomic background, and geographic characteristics (e.g., urbanicity level) across the US. We performed a GWAS of lifetime incidence of CP in the AoU v8 data, analyzing six ancestries and resulting in a total sample size of 291 081 individuals. We characterized genetic variants and genes associated with CP and estimated genetic correlations between CP and other physical health traits, psychiatric disorders, immune markers, and other traits of interest.

## Method

### Study Design

This study utilized data from the All of Us (AoU) research program^15^, a large-scale biobank based in the United States. Anyone residing in the United States who is 18 years of age or older is eligible to participate in the AoU research program, barring a few exceptions (e.g., in prison or suffering from a disability that renders them unable to provide consent). We used controlled-tier access data available from the version 8 release of the AoU Researcher Workbench. The version 8 release includes all individuals who had enrolled in the program by October 1^st^, 2023^16^. This study employed a cross-sectional case-control study design to investigate the genetic etiology of chronic pain.

### Study Participants

All participants with short-read whole genome sequencing (srWGS) and electronic health records (EHR) data available were considered for analysis (N = 317 964).

We excluded participants who were flagged by the AoU research team for relatedness based on a Hail kinship score of 0.1 or higher or failing genetic quality controls^17^ (N = 30 584), or whose self-reported sex at birth was not male or female (N = 3 334), resulting in a total analytic sample of 291 081 (*see* ***Table 1*** *for a detailed breakdown of sample sizes across ancestry and case-control status*).

**Table 1:** Breakdown of cases and controls for chronic pain across genetic ancestry.

| **Genetic Ancestry** | **Total N** | **N Cases** | **N Controls** |
| --- | --- | --- | --- |
| African (afr) | 56 794 | 25 344 | 31 450 |
| Admixed (amr) | 51 795 | 20 985 | 30 810 |
| East Asian (eas) | 6 658 | 1 824 | 4 834 |
| European (eur) | 171 011 | 87 239 | 83 772 |
| Middle Eastern (mid) | 1 120 | 481 | 639 |
| South Asian (sas) | 3 703 | 1 170 | 2 533 |
| **Total** | 291 081 | 137 043 | 154 038 |

### Definition of Lifetime Incidence of Chronic Pain

Using the cohort builder in the AoU researcher workbench, we defined a concept set of lifetime incidence of chronic pain (hereinafter referred to as chronic pain) based on SNOMED code 82423001. SNOMED codes are a medical coding scheme and are widely used by physicians to classify a patient’s condition in their EHR^18^. Chronic pain conditions ranged from chronic back pain to chronic postoperative pain (*refer to **Table S1***). We additionally included multiple medical conditions that are typically chronically painful (e.g., neuropathy; *see **Table S2***, which are usually but not necessarily painful) *only* if participants also self-reported having experienced pain greater than or equal to 3 (on a scale of 0-10) in the last 7 days (determined from item 1585747 in the ‘Overall Health’ survey; *see **Table S3***).

All individuals eligible for analysis (*see **Study Participants***) and with a condition in our chronic pain concept set were classified as cases (N = 137 043); all other individuals eligible for analysis but not in our chronic pain cohort were classified as controls (N = 154 038, *see* ***Table 1*** *for a detailed breakdown of case-control status across genetic ancestry*).

### Quality Control

All srWGS data were quality controlled by the AoU research team prior to analysis. For a detailed description of the quality controls the AoU research team implemented, please refer to the All of Us Genomic Data Quality Report^19^. Our team additionally filtered by minor allele frequency (MAF) > 1% to only include common single nucleotide polymorphisms (SNPs) and insertion/deletions in our analyses. While the number of single nucleotide polymorphisms (SNPs) tested varied across ancestry groups; on average, the number of SNPs tested was 15 016 293 (*see **Table S4***).

### Genome-wide association study of chronic pain

We used Plink2^20^ to conduct a genome-wide association study (GWAS) of chronic pain for each ancestry using a logistic regression model (plink2 command: --logistic ‘firth-fallback’). Across all analyses, we controlled for 10 within-ancestry common variant PCs, an individual’s age (derived as December/31/2024 minus date of birth), genotyping site (Broad Institute, University of Washington, or Baylor College of Medicine), and sex (male or female). To ensure the scales of continuous covariates did not vary too widely, we scaled age prior to conducting our analyses. Our genome-wide significance threshold was α = 5E-08, as is standard in GWAS.

Genetic ancestry was previously determined by the All of Us research team^17^. Within-ancestry common variant principal components (PCs) were generated using Plink2 following the Ricopili pipeline^21^ *(for scree plots for each ancestry, seeError! Reference source not found. **S1 - Figure S6***). To reduce potential confounds resulting from population stratification, we controlled for 10 ancestry-specific common PCs in all within-ancestry analyses.

### Cross-ancestry meta-analysis

We used mr-mega v. 0.2^22^ to meta-analyze our GWAS results across ancestries. mr-mega uses a meta-regression model and has previously been found to increase discovery power compared to traditional fixed- and random-effect meta-analysis and properly control for allelic heterogeneity between populations. We calculated the effective N for each single nucleotide polymorphism (SNP) as has been recently described^23^ and used these effective Ns as our sample size input for our meta-analysis. We additionally filtered for minor allele count (MAC) >= 10 and MAF >= 0.1 within mr-mega. After running the cross-ancestry meta-analysis, we further filtered for SNPs with an effective N >= 1000. We set our significance threshold for the cross-ancestry meta-analysis to α = 5E-08.

### FUMA annotation

We used FUMA v1.6.5^24^ to determine independent, genome-wide significant risk loci, annotate variants, and perform gene-wise analyses via MAGMA^25^. We defined “independent significant SNPs” as those which reached genome-wide significance (*p* < 5e-8) and were independent of each other at r^2^ < 0.6, and “lead SNPs” as those SNPs that were independent of each other at r^2^ < 0.1. “Genomic risk loci” were defined by merging linkage disequilibrium (LD) blocks of independent significant SNPs within a 250 kb distance. We used the appropriate ancestry-matched 1000 Genomes Phase 3 sample^26^ as the LD reference panel. We performed gene mapping in FUMA using positional mapping (based on ANNOVAR annotations), expression quantitative trait loci (eQTL) mapping (using GTEx V8^27^, CommonMind^28^, and Braineac^29^ data), and chromatin interaction mapping.

### Gene-based analyses

We performed MAGMA gene-set and gene property analyses (using GTEx V8 data) using the FUMA v1.6.5 platform.

### Genetic correlations

We used Popcorn v.1.0 ^30^ to estimate genetic correlations of CP across the three largest genetic ancestral populations included in this study (i.e., African, Admixed, and European). Popcorn provides two estimates: the genetic effect and the genetic impact. The genetic effect correlation (r_ge_) estimates the correlation between genetic effect sizes between two ancestries. Genetic impact correlation (r_gi_), on the other hand, accounts for population-specific allele differences and weighs population-specific common variants more heavily than rare variants within each population estimate. Within Popcorn, we estimated both the genetic effect (r_ge_) and genetic impact (r_gi_) correlations.

We used linkage disequilibrium score regression (LDSC^31,32^) to estimate the SNP-heritability of chronic pain in our European sample and the genetic correlations between chronic pain and 26 other phenotypes, including psychiatric disorders, behavioral traits, physical health conditions, immunological traits, and socioeconomic status-related phenotypes. Details on the individual GWAS used in genetic correlation analyses are provided in **Table S6**. In addition, we used LDSC to calculate the genetic correlation between chronic pain in our African sample and 6 other traits, with a focus on substance use and psychiatric disorders (*see **Table S8** for details*).

## Results

### Genomic loci associated with chronic pain

We identified one significant locus on chromosome 11 in the genome-wide association study (GWAS) in individuals of Admixed genetic ancestry, with the lead variant being an insertion/deletion mutation (indel) chr11:130015196:GA:G, rs5795684 (*see **Figure S7***, *p* = 1.13E-08). GWAS in the other five ancestries revealed no genome-wide significant variants *(see **Figure S8*** **– *Figure S11***). In the cross-ancestral meta-analysis, an indel, chr5:93838548:CA:C (SNP: rs1196962975) reached genome-wide significance *(p* = 4.53E-08*, see* ***Figure 1***). The CA allele at lead SNP rs1196962975 on chromosome 5 showed evidence of a protective effect in the AFR, AMR, and EUR ancestries (ORs = 0.90 - 0.97, ps = 3.95E-03 – 2.99E-05; *see* ***Figure 2***). rs1196962975 is an indel that resides within an intron of the *FAM172A*^33^ (gene aliases: *ARB2A* and *C5orf21*, ***Figure S13***). Past studies have found evidence linking this variant to changes in gene expression of *FAM172A* in immune-related cells: B-lymphocytes, T-lymphocytes, and other white blood cells^34,35^.

**Figure 1:**
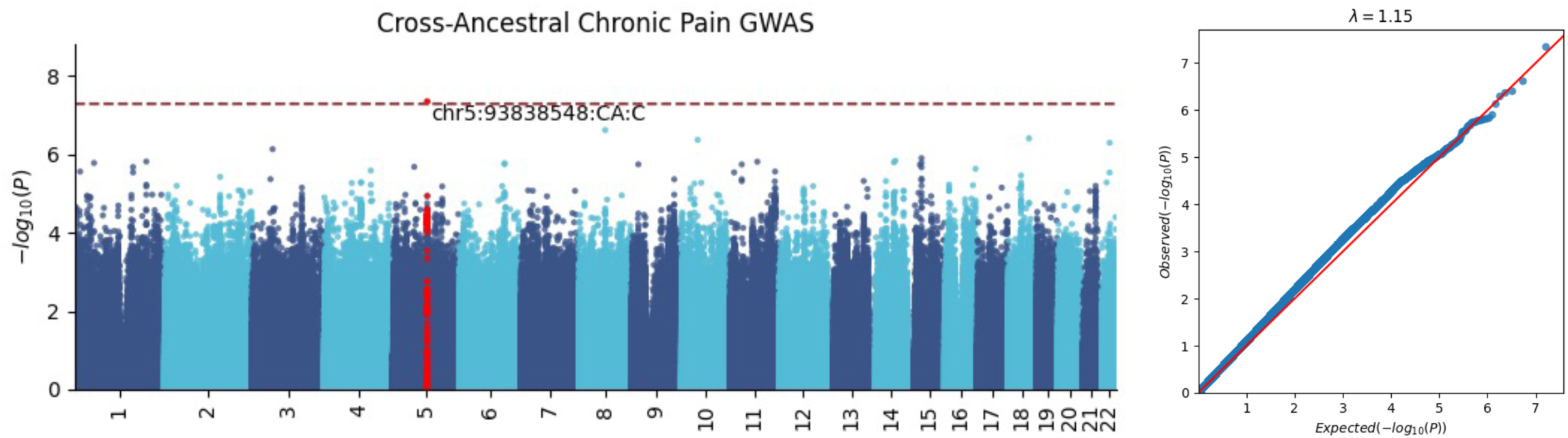
Cross-ancestral genome-wide association study of chronic pain.

**Figure 2:**
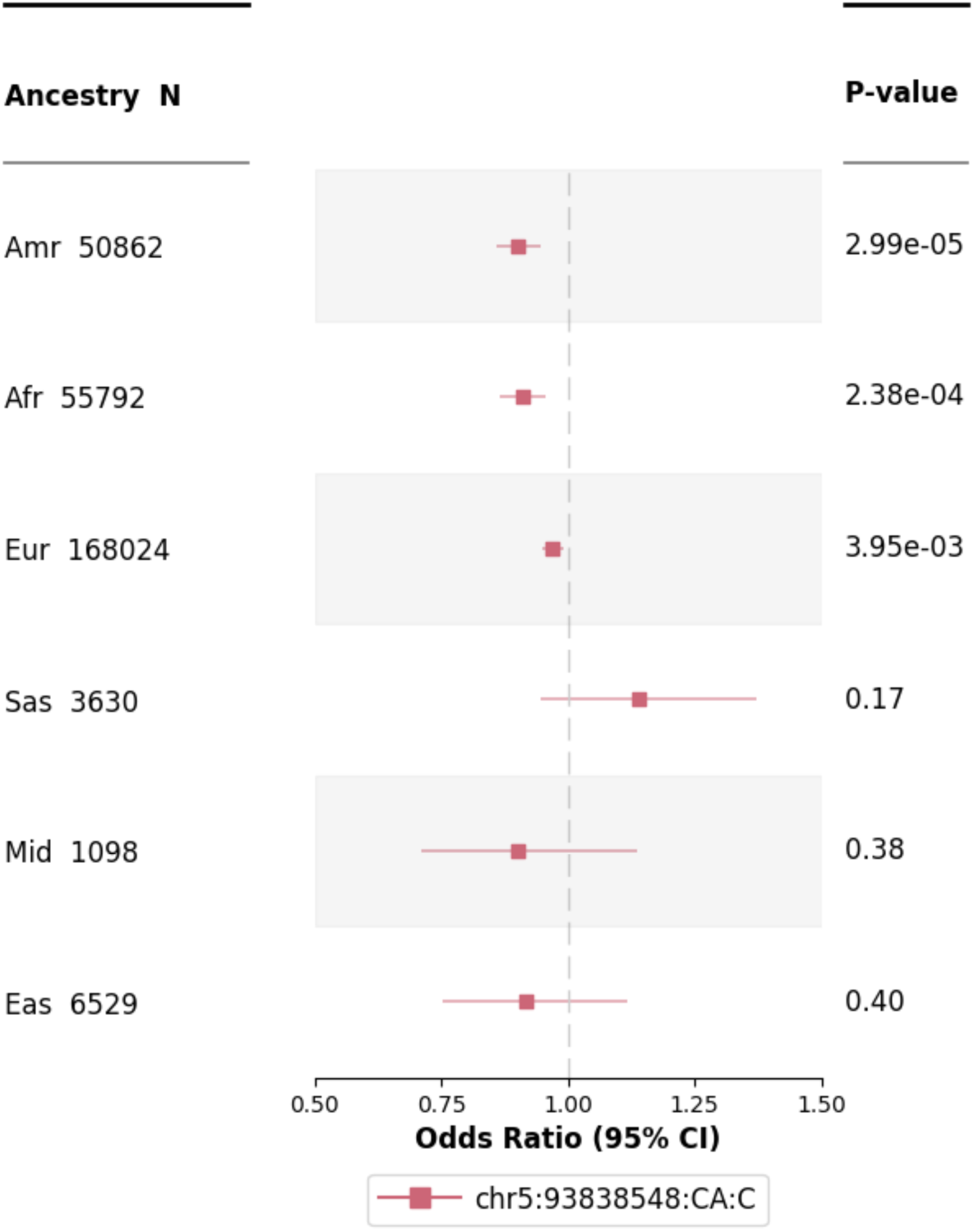
Forest plot of top locus (rs1196962975) across ancestries. Amr = Admixed; Afr = African; Eur = European; Sas = South Asian; Mid = Middle Eastern; Eas = East Asian.

The two variants identified in our study were not present in a previous GWAS of multisite chronic pain^11^ in a European ancestry subset of the UK Biobank. The variant that was significant in our GWAS of Admixed ancestry, rs5795684, is also an indel and might not have been included in past GWAS given most GWAS exclude indels from their analyses and is relatively uncommon (MAF_Euro_ = 0.049). The variant that was significant in our cross-ancestry meta-analysis, rs1196962975, is multiallelic, potentially explaining why it was not included in Johnston et al.’s study^11^. Of the 39 independent loci identified in that GWAS, 16 (41%) had nominally significant (*p* < 0.05) *p*-values in our European ancestry GWAS.

### Gene-based analyses

In the European ancestry data, there were no significant genes or gene-sets identified in the MAGMA analyses. However, the MAGMA gene-property analyses revealed significant enrichment in brain tissues (GTEx v8 30 general tissue types), specifically the nucleus accumbens, caudate, cerebellar hemisphere and cerebellum, hippocampus, and the amygdala (GTEx v8 53 specific tissue types; *see **Figure S14***).

### Cross-ancestral genetic correlations for chronic pain

Cross-ancestral genetic correlations for CP (*see* ***Table 2***) ranged from moderate (r_gi_ = 0.40 (SE = 0.37) to r_ge_ = 0.40 (SE = 0.40)) between the African and Admixed genetic ancestral groups and between the African and European ancestral groups (r_gi_ = 0.60 (SE = 0.29) to r_ge_ = 0.66 (SE = 0.34)) to high (r_gi_ = 0.78 (SE = 0.20) to r_ge_ = 0.76 (SE = 0.18)) between the Admixed and European subsets.

**Table 2:** Cross-ancestral genetic correlations for chronic pain. Afr = African; Amr = Admixed; Eur = European; rGE = genetic effect correlation; rGI = genetic impact correlation.

| Ancestry Comparison | $r_{GE}$ | $r_{GI}$ |
| --- | --- | --- |
| Afr-Amr | 0.40 (SE = 0.40) | 0.40 (SE = 0.37) |
| Amr-Eur | 0.76 (SE = 0.18) | 0.78 (SE = 0.20) |
| Afr-Eur | 0.66 (SE = 0.34) | 0.60 (SE = 0.29) |

### Genetic correlations between chronic pain and other traits

In the European ancestry subset, lifetime incidence of chronic pain was significantly genetically correlated with 24 out of the 26 traits tested (*see* Figure 3 *and **Table S7***, FDR-adjusted *p*-values < 0.05). First, our CP GWAS was strongly correlated with other previous studies of pain, including multisite chronic pain^11^ (r_g_ = 0.80, SE = 0.037, *p* = 3.14E-103), generalized pain^12^ (r_g_ = 0.91, SE = 0.043, *p* = 2.12E-99), and musculoskeletal pain^12^ (r_g_ = 0.46, SE = 0.047, *p* = 3.78E-23). Notably, the correlation with the generalized pain factor from Zorina-Lichtenwalter et al.^12^ was significantly stronger than the correlation with the musculoskeletal pain factor from that study (*p*_diff_ = 1.65e-13).

**Figure 3:**
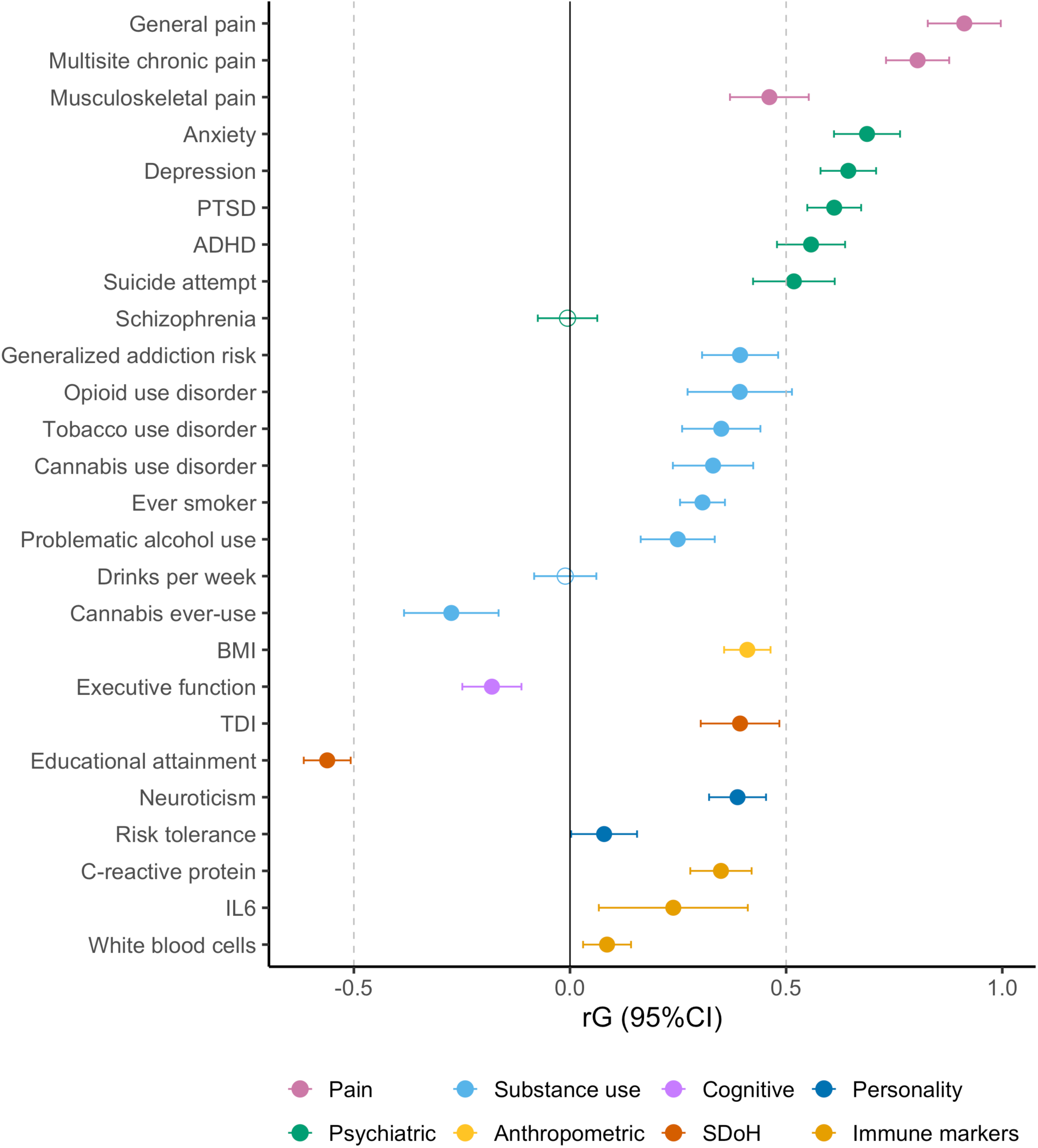
Genetic correlations with chronic pain. ADHD = attention deficit hyperactivity disorder; PTSD = post-traumatic stress disorder; BMI = body mass index; TDI = Townsend deprivation index; IL6 = interleukin-6.

CP was also strongly genetically correlated with multiple psychiatric disorders and traits, including ADHD (r_g_ = 0.56, SE = 0.040*, p* = 9.89E-44), depression (r_g_ = 0.64, SE = 0.033, *p* = 6.35E-86), anxiety (r_g_ = 0.69, SE = 0.039, *p* = 1.82E-69), post-traumatic stress disorder (PTSD, r_g_ = 0.61, SE = 0.032, *p* = 4.25E-82), and suicide attempts (r_g_ = 0.52, SE = 0.048, *p* = 5.93E-27). Schizophrenia, however, was not genetically correlated with CP (r_g_ = −0.0057, SE = 0.035, *p* = 0.87).

We also observed genetic correlations with substance use disorders, including generalized addiction risk (r_g_ = 0.39, SE = 0.045, *p* = 1.98E-18), opioid use disorder (r_g_ = 0.39, SE = 0.062, *p* = 1.84E-10), tobacco use disorder (r_g_ = 0.35, SE = 0.046, *p* = 3.54E-14), problematic alcohol use (r_g_ = 0.25, SE = 0.044, *p* = 1.15E-08), and cannabis use disorder (r_g_ = 0.33, SE = 0.047, *p* = 2.85E-12). CP was also genetically correlated with substance use behaviors (e.g., ever-smoking r_g_ = 0.31, SE = 0.027, *p* = 5.41E-31). However, CP was *negatively* genetically correlated with ever consuming cannabis (r_g_ = −0.27, SE = 0.056, *p* = 8.75E-07) and was not significantly correlated with drinks per week (r_g_ = −0.011, SE = 0.037, *p* = 0.76).

Our CP GWAS was also moderately genetically correlated with other cognitive and health-related traits, including a negative correlation with executive functioning (r_g_ = −0.18, SE = 0.035, *p* = 2.19E-07) and positive correlations with body mass index (r_g_ = 0.41, SE = 0.027, *p* = 1.55E-50) and several immune markers, including increased C-reactive protein serum levels (r_g_ = 0.35, SE = 0.036, *p* = 5.28E-22), interleukin-6 (IL-6, r_g_ = 0.24, SE = 0.088, *p* = 0.0066), and white blood cell count (r_g_ = 0.086, SE = 0.028, *p* = 0.0024).

Lastly, CP was also genetically correlated with several personality traits (neuroticism r_g_ = 0.39, SE = 0.034, *p* = 7.38E-31; increased risk tolerance r_g_ = 0.079, SE = 0.039, *p* = 0.042) and social determinants of health, with a positive correlation with the Townsend deprivation index, a measure of neighborhood deprivation, (r_g_ = 0.39, SE = 0.046, *p* = 2.11E-17) and a negative correlation with educational attainment (r_g_ = −0.56, SE = 0.028, *p* = 1.62E-91).

In the African subset, CP was not significantly genetically correlated with any of the traits tested; standard errors were substantially larger than in the European ancestry analyses (*see **Table S9***, FDR-adjusted *p*-values < 0.05). Though nonsignificant, the magnitude of some genetic correlation estimates were commensurate with those in the European subset. For example, the genetic correlation between CP and depression was high in the African subset (r_g_ = 0.80, SE = 0.52, *p* = 0.12), in line with the European estimate (r_g_ = 0.64, SE = 0.033). Similarly, the genetic correlation for PTSD in the African subset was estimated at r_g_ = 0.56 (SE = 0.34, *p* = 0.10), while the European estimate was r_g_ = 0.61.

## Discussion

Across the globe, lifetime incidence of chronic pain (CP) is exceedingly common^36,37^. Given its moderate heritability, there have been multiple genome-wide association studies (GWAS) of chronic pain and pain severity, but the majority have been confined to individuals of European ancestry. In this study, we conducted the most diverse genome-wide association study (GWAS) of broad CP to date using the All of Us Research Program biobank.

Error! Reference source not found.According to the Centers for Disease Control (CDC), in 2023, around 24.3% of U.S. adults were estimated to have chronic pain^38^. Notably, our prevalence was nearly double this estimate (∼ 47.08%, *see* ***Table 1***). We attribute our sample’s higher prevalence of chronic pain to multiple factors. First, we classified any individual who had ever had a lifetime incidence of chronic pain in our sample as endorsing chronic pain, whereas the CDC estimate is temporal (i.e., estimate of the number of adults actively experiencing chronic pain in 2023). Second, the All of Us Research program (AoU) is *reflective* of the U.S. population, but not necessarily *representative*^39^. AoU intentionally oversampled individuals of historically underrepresented groups in research, such as ethnic and sexual minorities^39^. These groups tend to disproportionately experience pain, including chronic pain^40^. Our study’s increased prevalence of lifetime incidence of chronic pain suggests that almost half of the U.S. population might experience CP sometime in their lifetime. Therefore, our study underscores the pervasiveness of CP and thereby the urgent need to further characterize and investigate medical treatments for CP using genetically and socioeconomically diverse samples.

Our genome-wide association study (GWAS) of CP identified two significant loci; one, on chromosome 11 (lead SNP: rs5795684, *p* = 1.13E-08), was significant only in the admixed genetic ancestry group. This locus resides within the *PRDM10* gene^41^, whose protein codes for transcription factors and positive regulatory domains containing C2H2-type zinc-fingers, which are involved in B cell differentiation and tumor suppression^42^. In addition, the gene homolog in mouse models has been implicated in central nervous system development^43^. The second significant locus resides on chromosome 5 and reached genome-wide significance after meta-analyzing across our six genetic ancestral groups (lead SNP: rs1196962975, *p* = 4.53E-08).

According to Open Targets Genetics, a database that integrates functional and biological databases and previous published studies, rs1196962975 is associated with changes in gene expression for genes involved in B and T immune cells^35^. Other studies^44^ have reported nominally (*p* < 0.001) significant associations between this variant and headache (*p* = 0.00070), back pain (*p* = 0.0014), had major operations (*p* = 0.0019), ever contemplated self-harm (*p* = 0.0024), and sarcoma/fibrosarcoma (*p* = 0.0033). This locus also lies within the *FAM172A* gene (gene aliases: *ARB2A* and *C5orf21)*. According to the human protein atlas^45^, *FAM172A* is involved in cell proliferation for various kinds of immune cells, including T cells, B cells, granulocytes, and monocytes, to name a few. Furthermore, Wei et al.^46^ found that removing the *FAM172A* promoter increased cell proliferation in mouse models of liver regeneration. This finding is in line with past research linking chronic pain and the immune system, especially in pain due to nerve injury or chronic inflammation^47^. For example, Vines et al.^48^ found that chronic back pain patients had significantly elevated levels of natural killer cell activity compared to controls. In short, our significant locus on chromosome 5 within the *FAM172A* gene is in line with previous studies and review articles^47,49,50^ that highlight the role of the immune system and immune function among mechanisms underlying the chronification of pain. This finding also exemplifies the benefit and importance of conducting multi-ancestral analyses to boost power (given that this locus was nonsignificant across individual ancestries) to identify genetic signals that converge across ancestral populations.

Our gene-based analyses in the European subset indicated that genomic signals were significantly more enriched in genes involved in brain areas including the nucleus accumbens, caudate, cerebellum, hippocampus, and the amygdala. These results are in line with substantial previous research implicating the roles of these brain regions in chronic pain^51^. For example, past research ^52^ has implicated the nucleus accumbens and the greater reward circuitry in the brain to the transition from acute to chronic pain. According to imaging studies, chronic lower back pain has been associated with impaired functionality in the hippocampus, caudate, and amygdala.

Moreover, recent research^53^ has identified cell and circuit changes in the amygdala’s central nucleus in the transition to chronic pain. In short, our gene and pathway analyses align with past research implicating the involvement of the nucleus accumbens, caudate, cerebellum, hippocampus, and the amygdala in chronic pain.

Our genetic correlation analyses revealed widespread genetic overlap between CP and a multitude of traits. First, our study was moderate to highly genetically correlated with past studies of multisite and generalized pain (r_g_ = [0.46 – 0.91]). In addition, our study was significantly less genetically correlated to a musculoskeletal pain factor (r_g_ = 0.46) compared to the generalized pain factor (r_g_ = 0.91, *p*_diff_ = 0.65E-13), further reflecting our broad definition of chronic pain in our sample. Our study was also highly genetically correlated with psychiatric traits such as depression (r_g_ = 0.64), PTSD (r_g_ = 0.61), and suicide attempts (r_g_ = 0.52), which have been extensively reported in the literature to be phenotypically comorbid with CP ^54,55^. In addition, CP was also genetically correlated with ADHD (r_g_ = 0.56); the link between CP and ADHD has been recently hypothesized to be mediated by neuroinflammation^56^, further supporting our top cross-ancestral significant loci (lead SNP: rs1196962975) located within a gene implicated in immune cell proliferation. Our study of CP was additionally modestly genetically correlated with substance use disorders (opioid use disorder r_g_ = 0.39, tobacco use disorder r_g_ = 0.35, cannabis use disorder r_g_ = 0.33, and generalized addiction risk r_g_ = 0.39).

These estimates are in line with Koller et al.’s^57^ reported genetic correlations between multisite chronic pain and substance use disorders (r_g_ = [0.20 – 0.37]). Our study was also genetically correlated with executive functioning (r_g_ = −0.18); a meta-analysis by Berryman et al.^58^ found that individuals with chronic pain exhibited small to medium cognitive impairments, and two studies reported that impairment in cognitive flexibility is associated with a higher risk of chronic postoperative pain^59,60^. Additionally, it is important to highlight that CP was negatively genetically correlated with educational attainment (r_g_ = −0.56) and positively genetically correlated with neighborhood disadvantage (r_g_ = 0.39). Previous demographic studies align with these findings. For example, Mullins et al.^61^ found that individuals with lower educational attainment were more likely to experience chronic pain in addition to lower pain management efficacy. Other studies^62^ have similarly found that neighborhood deprivation and neighborhood environmental factors impact chronic pain incidence and treatment. Lastly, CP was modestly genetically correlated to immune traits, including C-reactive protein (r_g_ = 0.35), IL6 (r_g_ = 0.24), and white blood cell count (r_g_ = 0.06). Like the top two loci associated with CP in this study, past studies have implicated the immune system in CP over the last two decades^63^. In fact, the role of the neuroimmune system on CP is emerging as an active and promising area of research^64^. In short, our study of CP is genetically correlated with past studies of chronic pain in addition to multiple sociobiological, neurobiological, immune, physical and psychiatric traits.

This study has several limitations. Our definition of chronic pain collapsed individuals with vastly different sources, locations, and intensities of pain. This broad phenotype – and the resulting heterogeneity - may have decreased our power to detect genetic loci and might explain our limited significant findings relative to other large GWAS of chronic pain phenotypes ^11,12^. In the future, dissecting subtypes of chronic pain might identify subtype-specific loci; additionally, using pain-matched controls might uncover additional common loci specifically associated with the chronification of pain. Lastly, our current study only examined common genetic variants (MAF > 1%). Future studies should expand analyses into rare variants to identify population-specific risk loci and test whether rare and common variants associated with CP co-act via similar or different biological pathways to influence CP.

In short, we conducted the first multi-ancestral genome-wide association study of broad chronic pain. Our results are aligned with established neurobiological and emerging immunological mechanisms underlying chronic pain. Lastly, our extensive genetic correlations between chronic pain and a variety of other socioeconomic factors and health traits aligns with previous research that conceptualizes chronic pain as a multifaceted and highly comorbid disease. Our high cross-ancestral genetic correlations suggest many genetic risk loci underlying chronic pain converge across ancestral populations and demonstrate the increased power to detect novel genetic loci by analyzing and pooling genetically diverse samples. Lifetime incidence of CP is high; it is increasingly urgent to further study CP to alleviate both human and economic suffering worldwide.

## Data Availability

The All of Us Research Program data are available to registered and approved All of Us researchers. Genetic data requires controlled tier access; researchers can register for access through their institutions. Summary statistics resulting from this study will be deposited to the GWAS Catalog upon publication. Code for this project will be available upon publication on the All of Us workbench as a public workspace and on GitHub.

## Acknowledgements

The All of Us Research Program is supported by the National Institutes of Health, Office of the Director: Regional Medical Centers: 1 OT2 OD026549; 1 OT2 OD026554; 1 OT2 OD026557; 1 OT2 OD026556; 1 OT2 OD026550; 1 OT2 OD 026552; 1 OT2 OD026553; 1 OT2 OD026548; 1 OT2 OD026551; 1 OT2 OD026555; IAA #: AOD 16037; Federally Qualified Health Centers: HHSN 263201600085U; Data and Research Center: 5 U2C OD023196; Biobank: 1 U24 OD023121; The Participant Center: U24 OD023176; Participant Technology Systems Center: 1 U24 OD023163; Communications and Engagement: 3 OT2 OD023205; 3 OT2 OD023206; and Community Partners: 1 OT2 OD025277; 3 OT2 OD025315; 1 OT2 OD025337; 1 OT2 OD025276. In addition, the All of Us Research Program would not be possible without the partnership of its participants. This manuscript is the result of funding in whole or in part by the National Institutes of Health (NIH). It is subject to the NIH Public Access Policy. Through acceptance of this federal funding, NIH has been given a right to make this manuscript publicly available in PubMed Central upon the Official Date of Publication, as defined by NIH. This work utilized the High Throughput Computing Facility at the Center for Genome Sciences and Systems Biology at Washington University in St. Louis.

## Funding

This work was supported by the National Institute for Drug Abuse [grant numbers R03DA059747, K01DA051759, T32DA007261, and R01DA054869]; the National Institute of Nursing Research [grant number R90NR021799]; the National Institute on Alcohol Abuse and Alcoholism [grant number K01AA030083]; and the National Institute on Neurological Disorders and Stroke [grant number RM1NS135283].

## Disclosures

PNRV, JVB, NS, ASH, AA, and ECJ report no conflicts of interest. SH discloses personal fees from Vertex.

