## Supplementary Information for "Multi-ancestral genome-wide study of chronic pain reveals widespread genetic correlations with mental and physical health traits"

### Supplementary Tables and Figures

***Table S 1: List of pain conditions included using only SNOMED code 82423001 of chronic pain.***

| Condition | Concept ID in Data Browser |
|---|---|
| Bilateral chronic pain of upper limbs | 35615072 |
| Chronic back pain | 4046660 |
| Chronic chest pain | 762941 |
| Chronic interstitial cystitis | 75863 |
| Chronic musculoskeletal pain | 42538688 |
| Chronic neck pain | 43530622 |
| Chronic neuropathic pain | 42536909 |
| Chronic pain | 436096 |
| Chronic pain following left total knee arthroplasty | 37108780 |
| Chronic pain following right total knee arthroplasty | 37117142 |
| Chronic pain in face | 4196899 |
| Chronic pain of left foot | 37108974 |
| Chronic pain of left upper limb | 37108941 |
| Chronic pain of right foot | 37108973 |
| Chronic pain of right upper limb | 37108940 |
| Chronic pain syndrome | 440704 |
| Chronic postoperative pain | 43531612 |
| Complex regional pain syndrome | 4134577 |

***Table S 2: Characteristically painful medical conditions that were additionally included in our study's definition of chronic pain.***

| Condition | Concept ID in Data Browser |
|---|---|
| Neuralgia | 373852 |
| Peripheral neuralgia | 4101480 |
| Otalgia | 380733 |
| Fibromyalgia | 40405599 |
| Metatarsalgia | 4002956 |
| Cervico-occipital neuralgia | 4216083 |
| Trigeminal neuralgia | 379801 |

| | |
|---|---|
| Trigeminal autonomic cephalalgia | 42709942 |
| Referred otalgia | 375825 |
| Otogenic otalgia | 374372 |
| Postherpetic neuralgia | 4071164 |
| Morton's metatarsalgia | 4107377 |
| Post-herpetic trigeminal neuralgia | 381504 |
| Scapulalgia | 4049524 |
| Erythromelalgia | 134380 |
| Bilateral metatarsalgia | 760924 |
| Tenalgia | 4068332 |
| Supraorbital neuralgia | 4068401 |
| Glossopharyngeal neuralgia | 380100 |
| Right trigeminal neuralgia | 760909 |
| Left trigeminal neuralgia | 760908 |
| Polymyalgia | 4319324 |
| Intercostal neuralgia | 4090559 |
| Facial neuralgia | 43530746 |
| Intercostal myalgia | 4318397 |
| Pudendal neuralgia | 4176933 |
| Cyclical mastalgia | 4034226 |
| Non-cyclical mastalgia | 4130004 |
| Brachial plexus neuralgia | 4297883 |
| Genitofemoral nerve neuralgia | 4083775 |
| Cranial neuralgia | 4045968 |
| Iliohypogastric nerve neuralgia | 4090562 |
| Ilioinguinal nerve neuralgia | 4083291 |
| Perineal neuralgia | 4090561 |
| Post-surgery obturator neuralgia | 37017552 |
| Viral myalgia | 4344370 |
| Dysuria | 197684 |
| Generalized abdominal pain | 197988 |
| Disorder characterized by pain | 4163285 |
| Intermittent pain | 4200298 |
| Chronic thoracic back pain | 43530661 |
| Phantom limb syndrome with pain | 45773181 |
| Chronic ankle pain | 45763561 |
| Chronic sacroiliac joint pain | 37204166 |
| Chronic abdominal pain | 4008102 |
| Peripheral neuropathic pain | 4089698 |

| | |
|---|---|
| Recurrent abdominal pain | 4258543 |
| Disorder characterized by back pain | 4163285 |
| Chronic intractable pain | 4168685 |
| Uncontrolled pain | 46270108 |
| Chronic urinary bladder pain | 37209550 |
| Persistent testicular pain | 4107202 |
| Chronic central neuropathic pain | 42538797 |
| Chronic vaginal pain | 4142567 |
| Generalized chronic body pains | 4010338 |
| Generalized acute body pains | 4010834 |
| Generalized aches and pains | 438867 |
| Endometriosis (clinical) | 433527 |
| Polyalgia | 4344019 |
| Chronic tension-type headache | 377853 |
| Chronic post-traumatic headache | 377546 |
| Chronic cluster headache | 378145 |
| Chronic daily headache | 36684905 |
| Intractable chronic tension headache | 42535410 |
| Chronic mixed headache syndrome | 4318560 |
| Chronic post-concussion headache | 44783586 |
| Migraine | 318736 |
| Chronic intractable migraine without aura | 43530652 |
| Menstrual migraine | 376394 |
| Cluster headache | 381278 |

***Table S 3: Neuropathic conditions that were included in our study's definition of chronic pain only if the participant additionally self-reported having experienced significant pain in the last week.***

| **Condition** | **Concept ID in Data Browser** |
|---|---|
| Neuropathy due to diabetes mellitus | 4044391 |
| Peripheral neuropathy due to type 2 diabetes mellitus | 37016354 |
| Neuropathy | 4301699 |

***Table S 4: Number of SNPs tested across ancestries.***

| **Ancestry** | **N SNPs Tested** | **N SNPs Converged** |
|---|---|---|
| African (afr) | 21 299 885 | 20 833 835 |
| Admixed (amr) | 14 883 306 | 13 967 282 |
| East Asian (eas) | 12 093 324 | 11 114 480 |
| European (eur) | 12 976 437 | 12 495 520 |
| Middle Eastern (mid) | 15 314 427 | 13 744 146 |
| South Asian (sas) | 13 530 379 | 12 768 417 |
| **Average** | 15 016 293 | 14 153 946.6667 |

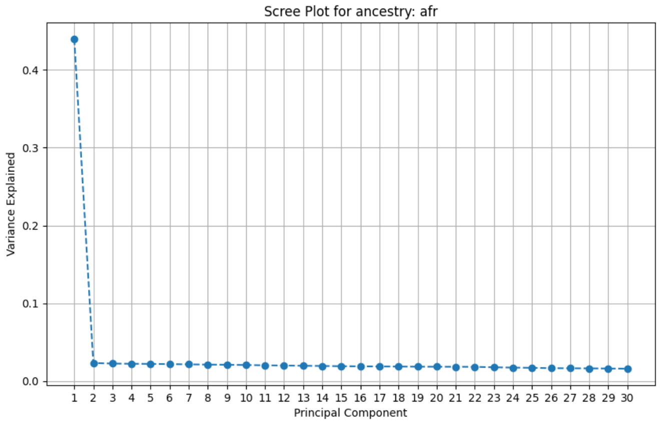


***Figure S 1: Variance explained of common variant principal components in individuals of African genetic ancestry.***

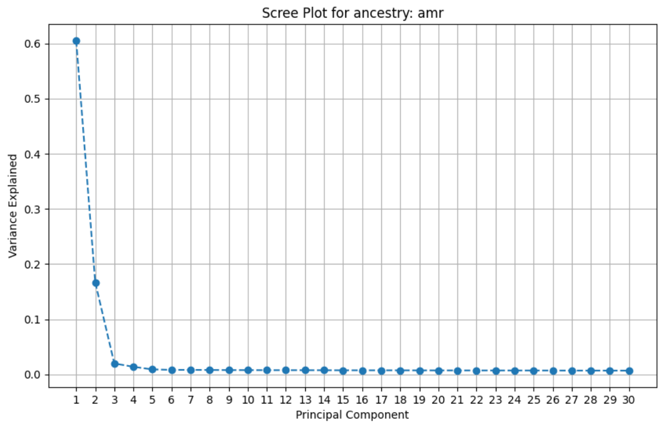


***Figure S 2: Variance explained by common PCs for individuals of Admixed genetic ancestry.***

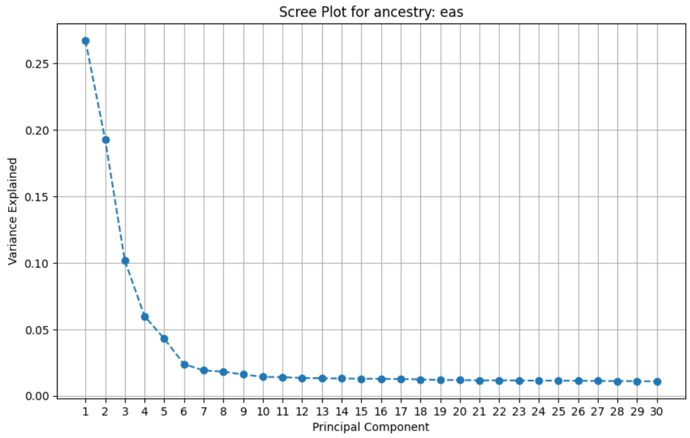


***Figure S 3: Variance explained by within-ancestry common PCs in the East Asian population.***

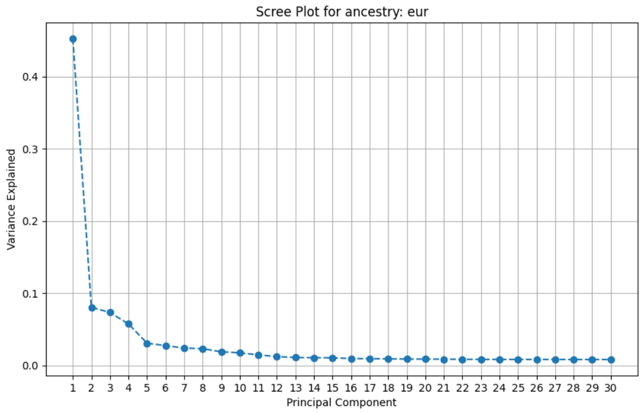


***Figure S 4: Variance explained by within-ancestry common PCs in the European population.***

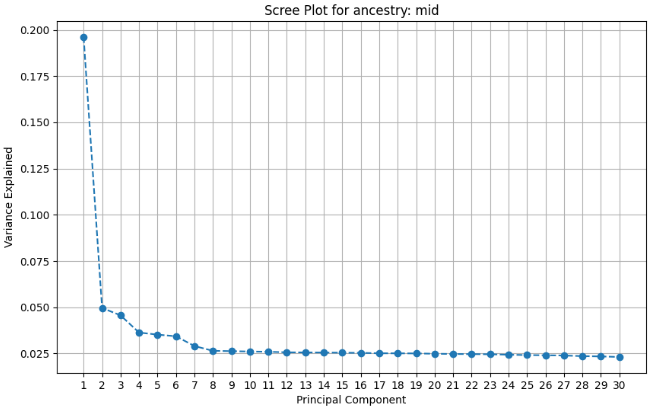


***Figure S 5: Variance explained by within-ancestry common PCs in the Middle Eastern ancestral population.***

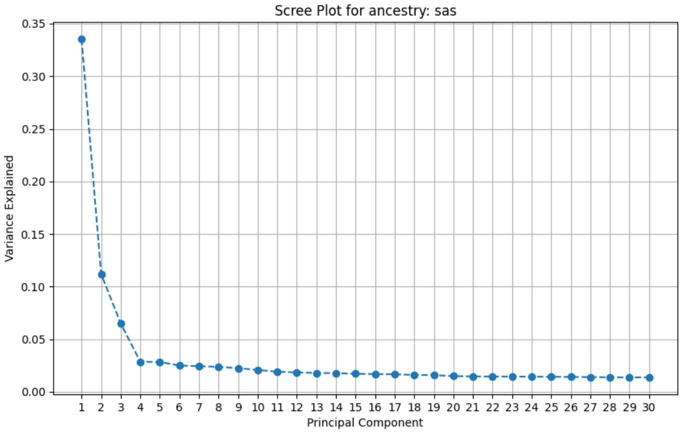


***Figure S 6: Variance explained by within-ancestry common PCs in the South Asian ancestral population.***

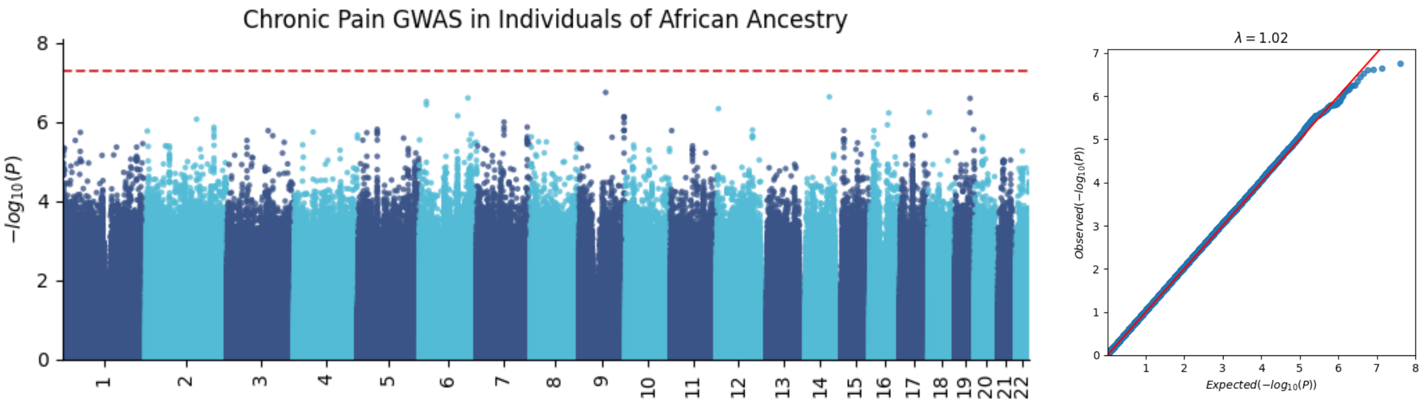


***Figure S 7: Genome-wide association study of chronic pain in individuals of Admixed genetic ancestry.***

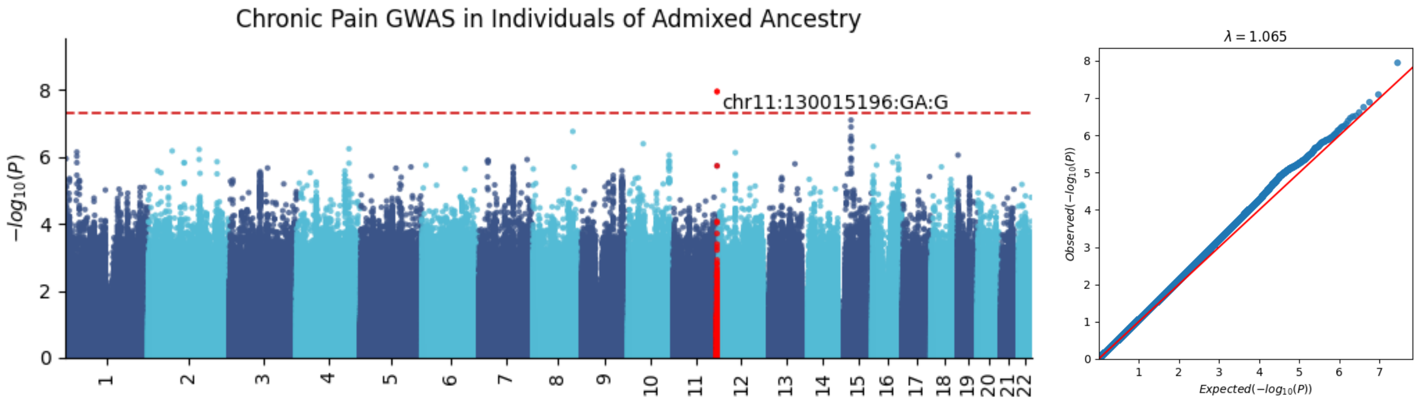


***Figure S 8: Genome-wide association study of chronic pain in individuals of African genetic ancestry.***

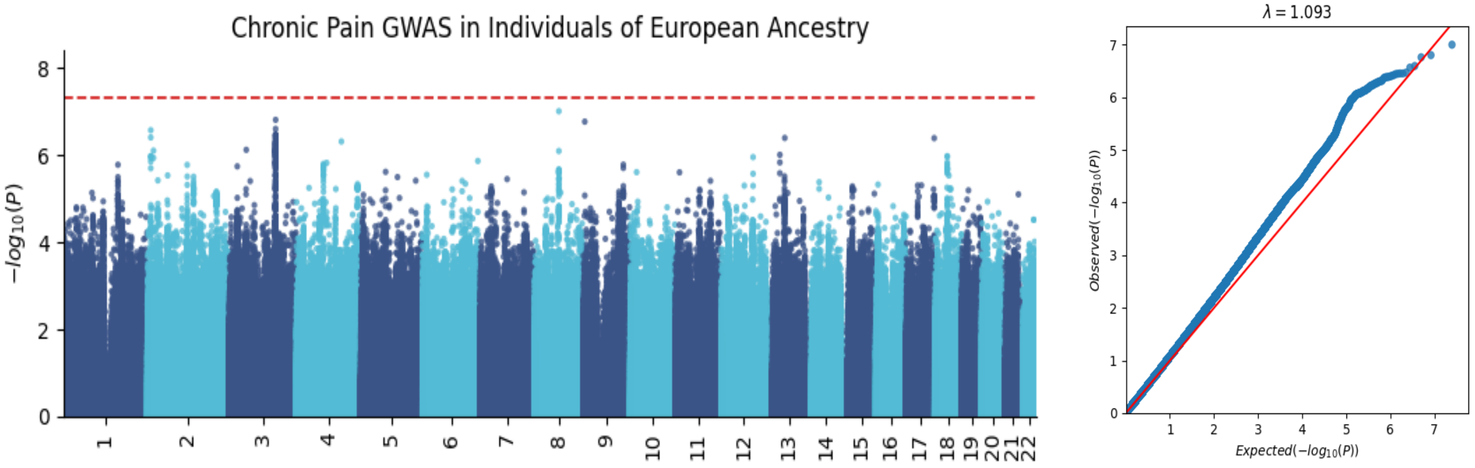


***Figure S 9: Genome-wide association study of chronic pain in individuals of European genetic ancestry.***

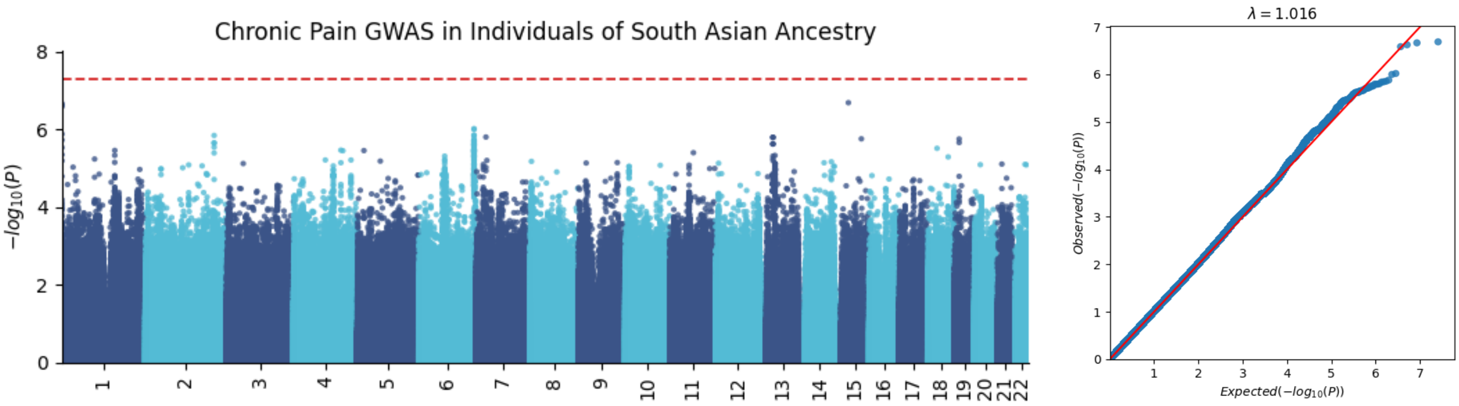


***Figure S 12: Genome-wide association study of chronic pain in individuals of South Asian genetic ancestry.***

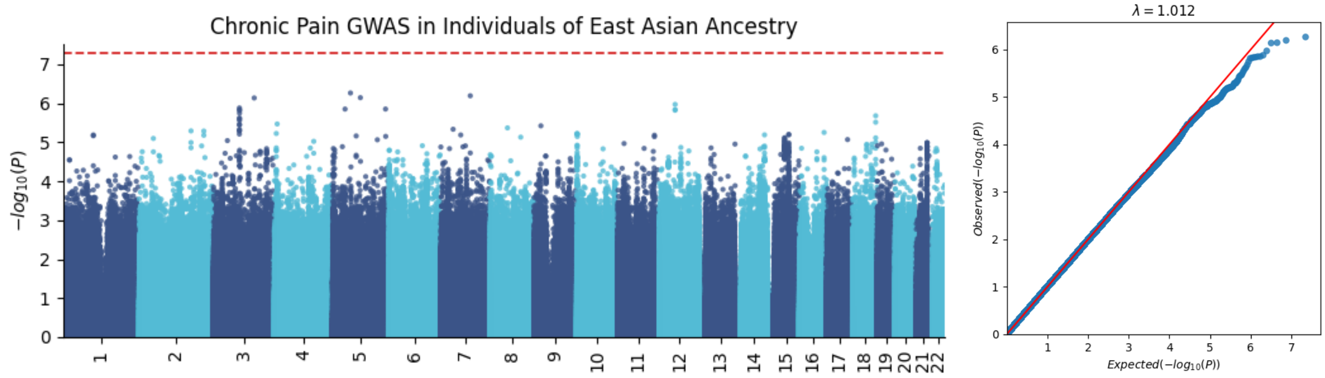


***Figure S 11: Genome-wide association study of chronic pain in individuals of East Asian genetic ancestry.***

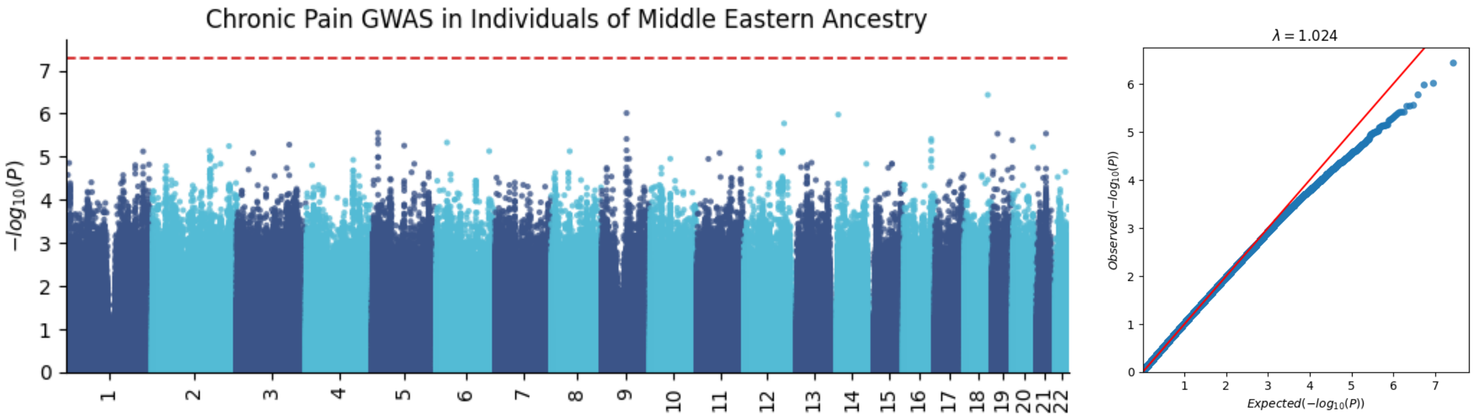


***Figure S 10: Genome-wide association study of chronic pain in individuals of Middle Eastern genetic ancestry.***

***Table S 5: P-values from the top 39 loci found by Johnston et al. 2019 in our study.***

| **Genomic Locus** | **Chromosome** | **SNP** | **SNP *p*-value in EUR ancestry in current study** | **SNP *p*-value in Johnston et al.** |
|---|---|---|---|---|
| 1 | 1 | rs10888692 | 5.53E-02 | 5.30E-09 |
| 2 | 1 | rs35311109 | 7.99E-03 | 2.00E-09 |
| 3 | 1 | rs197422 | 0.138 | 9.20E-12 |
| 4 | 1 | rs12033257 | 5.35E-03 | 5.30E-09 |
| 5 | 2 | rs4852567 | 0.412285 | 4.30E-08 |
| 6 | 3 | rs7628207 | 2.75E-04 | 8.40E-10 |
| 7 | 3 | rs28428925 | 0.154949 | 1.40E-09 |
| 8 | 3 | rs6770476 | 3.22E-04 | 9.40E-09 |
| 9 | 4 | rs34811474 | 0.480447 | 2.70E-11 |
| 10 | 4 | rs13135092 | 3.07E-02 | 1.50E-13 |
| 11 | 4 | rs13136239 | 1.45E-03 | 3.60E-08 |
| 12 | 5 | rs6869446 | 0.234993 | 9.50E-09 |
| 13 | 5 | rs1976423 | 0.569657 | 8.20E-09 |
| 14 | 5 | rs17474406 | 0.784437 | 2.40E-08 |
| 15 | 5 | rs1946247 | 0.141197 | 4.90E-08 |
| 16 | 6 | rs11751591 | 0.15352 | 2.70E-10 |
| 17 | 6 | rs6907508 | 1.57E-02 | 1.10E-08 |
| 18 | 6 | rs6926377 | 0.951863 | 7.90E-09 |
| 19 | 7 | rs10259354 | 0.604888 | 3.00E-08 |
| 20 | 7 | rs7798894 | 4.33E-04 | 1.60E-08 |
| 21 | 7 | rs6966540 | 0.378427 | 3.30E-08 |
| 22 | 7 | rs12537376 | 2.19E-03 | 1.70E-09 |
| 23 | 8 | rs11786084 | 4.18E-02 | 2.30E-08 |
| 24 | 9 | rs10992729 | 9.47E-02 | 1.10E-09 |
| 25 | 9 | rs6478241 | 0.941451 | 3.10E-09 |
| 26 | 9 | 9:140251458:G:A | 3.73105E-03 | 5.30E-14 |
| 27 | 10 | rs2183271 | 0.272218 | 3.10E-08 |
| 28 | 10 | rs11599236 | 0.124082 | 3.30E-08 |
| 29 | 10 | rs12765185 | 4.77E-02 | 3.90E-08 |
| 30 | 11 | rs61883178 | 7.81E-02 | 2.00E-10 |
| 31 | 13 | rs1443914 | 0.133887 | 2.80E-11 |
| 32 | 14 | rs12435797 | 0.352742 | 3.70E-08 |
| 33 | 14 | rs2006281 | 0.40899 | 3.40E-08 |
| 34 | 15 | rs2386584 | 3.49E-02 | 2.80E-11 |

| | | | | |
|---|---|---|---|---|
| 35 | 16 | rs285026 | 0.189704 | 1.90E-08 |
| 36 | 17 | rs11871043 | 4.10E-02 | 1.70E-09 |
| 37 | 17 | rs11079993 | 5.93E-04 | 5.70E-12 |
| 38 | 18 | rs62098013 | 4.96E-02 | 4.00E-11 |
| 39 | 20 | rs2424248 | 5.86E-02 | 3.70E-10 |

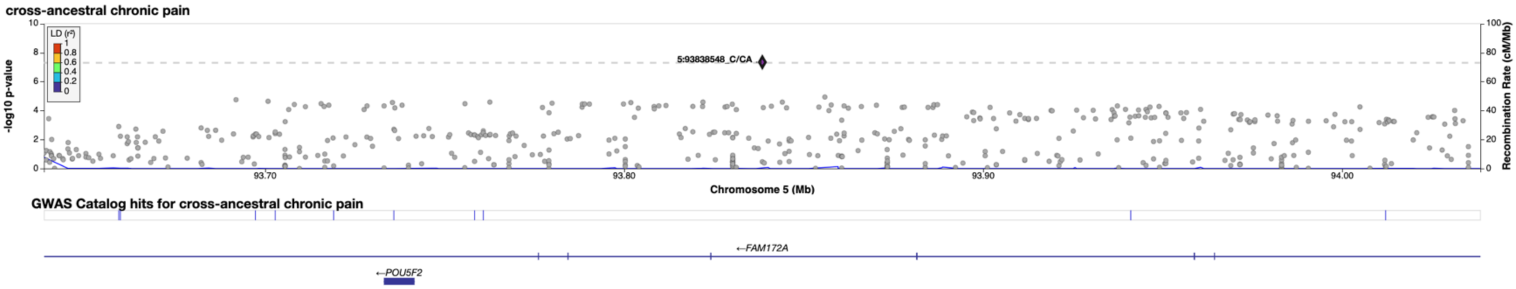


*Figure S 13: Locus Zoom plot of top cross-ancestral loci*

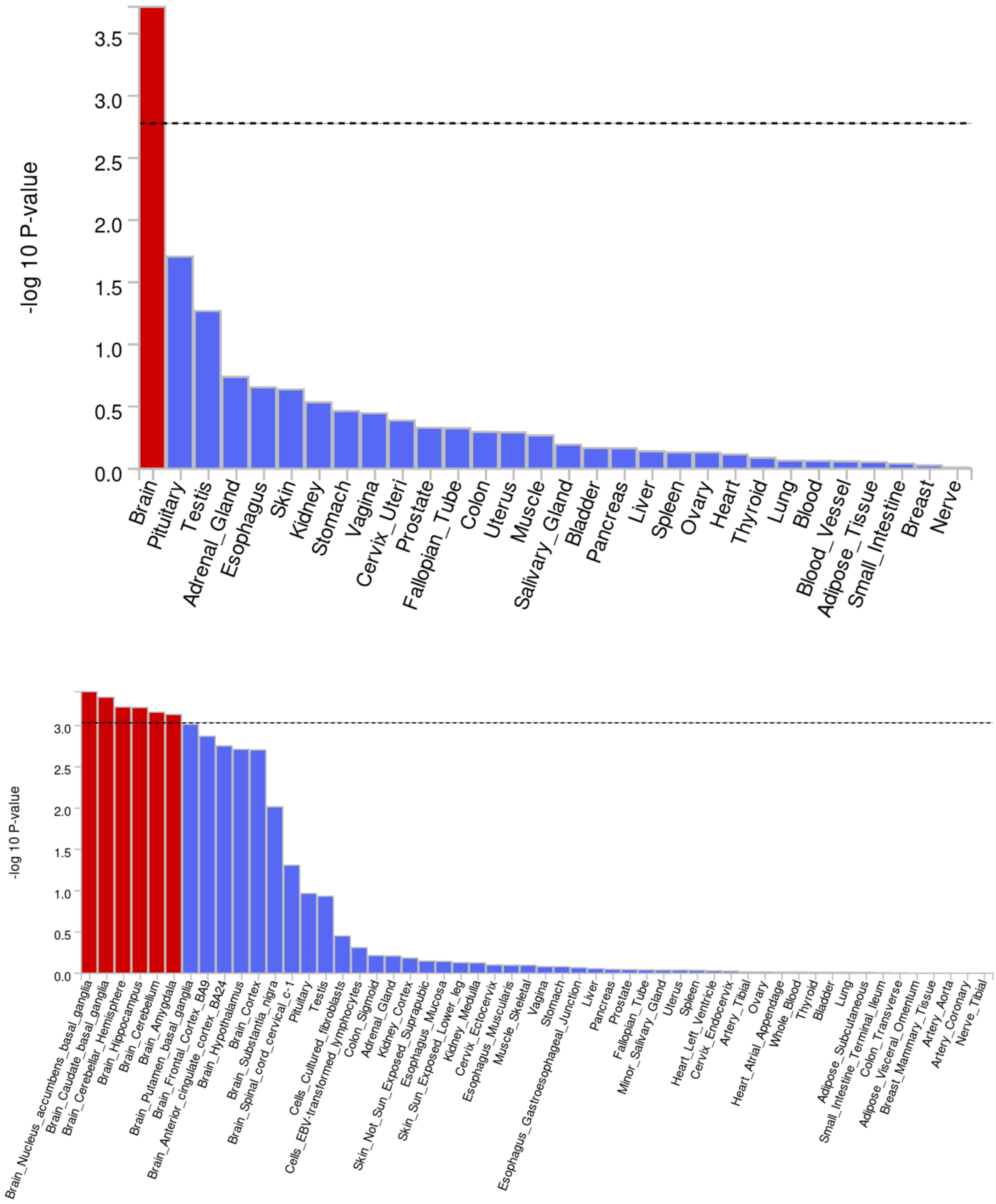


***Figure S 14: MAGMA gene-property analyses.***

Top: Comparing 30 general tissues, we found a significant enrichment in the brain.
Bottom: Using 53 organ tissues, we found significant enrichment in brain tissues, including the nucleus accumbens, basal ganglia, hippocampus, cerebellum, and amygdala.

***Table S 6: Studies included in genetic correlation analyses with the European sample.***

ADHD = attention deficit hyperactivity disorder; PTSD = post-traumatic stress disorder; BMI = body mass index; TDI = Townsend deprivation index; IL6 = Interleukin-6

| **Trait** | **Study PMID** |
|---|---|
| Multisite Chronic Pain | 31194737 |
| General Pain | 37219871 |
| Musculoskeletal Pain | 37219871 |
| ADHD | 36702997 |
| Anxiety | 39294497 |
| Depression | 39814019 |
| PTSD | 38637617 |
| Schizophrenia | 35396580 |
| Suicide Attempt | 37777856 |
| Generalized Addiction Risk | 37250466 |
| Opioid Use Disorder | 35879402 |
| Tobacco Use Disorder | 38632388 |
| Cannabis Use Disorder | 37985822 |
| Ever Smoker | 36477530 |
| Problematic Alcohol Use | 38062264 |
| Drinks per Week | 36477530 |
| Cannabis Ever Use | 30150663 |
| BMI | 30124842 |
| Executive Function | 36150907 |
| TDI | No PMID. Link: http://www.nealelab.is/uk-biobank/ |
| Educational Attainment | 35361970 |
| Neuroticism | 29942085 |
| Risk Tolerance | 30643258 |
| C-reactive Protein | 35459240 |
| IL6 | 33517400 |
| White Blood Cell Count | 32888493 |

***Table S 7: Genetic correlations between our study's European sample of chronic pain and GWAS of other traits.***

rg = genetic correlation; SE = standard error; ADHD = attention deficit hyperactivity disorder; PTSD = post-traumatic stress disorder; BMI = body mass index; TDI = Townsend deprivation index; SDoH = social determinants of health; IL6 = Interleukin-6

| **Trait** | **Domain** | **rg** | **SE** | ***p*-value** |
|---|---|---|---|---|
| Multisite Chronic Pain | Pain | 0.8041 | 0.0373 | 3.14E-103 |
| General Pain | Pain | 0.9121 | 0.0431 | 2.12E-99 |
| Musculoskeletal Pain | Pain | 0.4612 | 0.0465 | 3.78E-23 |
| ADHD | Psychiatric | 0.5576 | 0.0402 | 9.89E-44 |
| Anxiety | Psychiatric | 0.6871 | 0.039 | 1.82E-69 |
| Depression | Psychiatric | 0.6438 | 0.0328 | 6.35E-86 |
| PTSD | Psychiatric | 0.6112 | 0.0318 | 4.25E-82 |
| Schizophrenia | Psychiatric | -0.0057 | 0.0351 | 0.8711 |
| Suicide Attempt | Psychiatric | 0.5179 | 0.0482 | 5.93E-27 |
| Generalized Addiction Risk | Substance use and/or disorders | 0.3935 | 0.0449 | 1.98E-18 |
| Opioid Use Disorder | Substance use and/or disorders | 0.3927 | 0.0616 | 1.84E-10 |
| Tobacco Use Disorder | Substance use and/or disorders | 0.3499 | 0.0462 | 3.54E-14 |
| Cannabis Use Disorder | Substance use and/or disorders | 0.3309 | 0.0474 | 2.85E-12 |
| Ever Smoker | Substance use and/or disorders | 0.3065 | 0.0265 | 5.41E-31 |
| Problematic Alcohol Use | Substance use and/or disorders | 0.2492 | 0.0437 | 1.15E-08 |
| Drinks per Week | Substance use and/or disorders | -0.011 | 0.0367 | 0.7641 |
| Cannabis Ever Use | Substance use and/or disorders | -0.2746 | 0.0558 | 8.75E-07 |
| BMI | Anthropometric | 0.4102 | 0.0274 | 1.55E-50 |
| Executive Function | Cognitive | -0.1807 | 0.0349 | 2.19E-07 |
| TDI | SDoH | 0.3934 | 0.0464 | 2.11E-17 |
| Educational Attainment | SDoH | -0.5616 | 0.0277 | 1.62E-91 |
| Neuroticism | Personality | 0.3876 | 0.0336 | 7.38E-31 |
| Risk Tolerance | Personality | 0.0789 | 0.0389 | 0.0424 |
| C-reactive Protein | Immune | 0.3492 | 0.0362 | 5.28E-22 |

| | | | | |
|---|---|---|---|---|
| IL6 | Immune | 0.239 | 0.0879 | 0.0066 |
| White Blood Cell Count | Immune | 0.0857 | 0.0283 | 0.0024 |

***Table S 8: Studies included in genetic correlation analyses with our study's African sample.***

PTSD = post-traumatic stress disorder.

| **Trait** | **PMID** |
|---|---|
| Anxiety | 31906708 |
| Depression | 34045744 |
| PTSD | 38637617 |
| Cannabis Use Disorder | 37985822 |
| Ever Smoker | 36477530 |
| Drinks per Week | 36477530 |

***Table S 9: Genetic correlations between our study's African sample of chronic pain and GWAS of other traits.***

PTSD = post-traumatic stress disorder.

| **Trait** | **Domain** | **rg** | **SE** | ***p*-value** |
|---|---|---|---|---|
| Anxiety | Psychiatric | -0.0395 | 0.4247 | 0.9259 |
| Depression | Psychiatric | 0.8043 | 0.5186 | 0.1209 |
| PTSD | Psychiatric | 0.5633 | 0.3449 | 0.1024 |
| Cannabis Use Disorder | Substance use and/or disorders | -0.08032 | 0.3006 | 0.7819 |
| Ever Smoker | Substance use and/or disorders | 0.1284 | 0.2956 | 0.6639 |
| Drinks per Week | Substance use and/or disorders | 0.1306 | 0.6133 | 0.8314 |
